# Use of Public Data to Describe COVID-19 Contact Tracing in China during January 20–February 29, 2020

**DOI:** 10.1101/2020.12.04.20243972

**Authors:** Emilio Dirlikov, Suizan Zhou, Lifeng Han, Zhijun Li, Ling Hao, Alexander J. Millman, Barbara Marston

## Abstract

**Objective:** Although contact tracing is generally not used to control influenza pandemics, China and several countries in the Western Pacific Region employed contact tracing as part of COVID-19 response activities. To improve understanding on the use of contact tracing for COVID-19 emergency public health response activities, we describe reported COVID-19 contacts traced and quarantined in China and a proxy for number of reported contacts traced per reported case.

**Methods:** We abstracted publicly available online aggregate data reported from China’s National Health Commission and provincial health commissions’ COVID-19 daily situational reports for January 20–February 29, 2020. The number of new contacts traced by report date was computed as the difference between total contacts traced on consecutive reports. A proxy for the number of contacts traced per case was computed as the number of new contacts traced divided by the number of new cases.

**Results:** During January 20–February 29, 2020, China reported 80,968 new COVID-19 cases (Hubei Province = 67,608 [83%]), and 659,899 contacts traced (Hubei Province = 265,617 [40%]). Non-Hubei provinces reported more contacts traced per case than Hubei Province; this difference increased over time.

**Discussion:** Along with other NPI used in China, contact tracing likely contributed to reducing SARS-CoV-2 transmission by quarantining a large number of potentially infected contacts. Despite reporting only 15% of total cases, non-Hubei provinces had 1.5 times more reported contacts traced compared to Hubei Province. Contract tracing may have been more complete in areas and periods with lower case counts.

## Introduction

Coronavirus disease 2019 (COVID-19) is a respiratory illness caused by severe acute respiratory syndrome coronavirus 2 (SARS-CoV-2), first identified in December 2019 in Hubei Province, China (1). On January 20, 2020, China National Health Commission (NHC) reported evidence of person-to-person transmission and began reporting daily COVID-19 situational reports. By January 31, 2020, ≥1 case had been reported from each of mainland China’s 31 provincial-level administrative units. A total of 82,875 cases were reported by May 1, of which 96% had been reported by February 29 (2).

Non-pharmaceutical interventions (NPIs) for respiratory viruses aim to reduce transmission through individual or community actions, by preventing exposures. (3). China implemented COVID-19 contact tracing with quarantine as part of a comprehensive COVID-19 prevention and control strategy, which also includes mask use, emphasis of hand hygiene, enforced social distancing, and travel restrictions (8, 9).

Contact tracing is generally not used to control influenza pandemics because of short generation time for cases and resource constraints (4). However, China and several countries employed contact tracing as part of COVID-19 response activities (5-7). China’s contact tracing strategy aimed to identify and quarantine exposed individuals to prevent additional disease transmission. On January 20, 2020, China designated COVID-19 a notifiable disease and updated the “Frontier Health and Quarantine Law” to allow quarantine of contacts (10). National guidelines on epidemiologic investigations and the management of contacts were issued and updated several times, and delegated responsibility for contact tracing to the local level (11, 12). The latest guidelines define contacts as: “anyone who may have had contact with a case through a range of circumstances or activities including being family members, relatives, friends, colleagues, classmates, health care workers, and services personnel” (12).

Here, we describe the reported COVID-19 contacts traced and a proxy for number of reported contacts traced per reported case in China during January 20–February 29, 2020.

## Methods

We abstracted publicly available online aggregate data reported from NHC and provincial health commissions’ COVID-19 daily situational reports (Technical Appendix). For reports during January 20–February 29 (epidemiologic weeks 4–9), we collected daily reported data on: newly reported cases and total contacts traced and placed under medical observation. Data were reviewed for abstraction errors. Provinces with >95% data completeness were included.

The number of new contacts traced by report date was computed as the difference between total contacts traced on consecutive reports. A proxy for the number of contacts traced per case was computed as the number of new contacts traced divided by the number of new cases. These calculations were performed by epidemiologic week.

In additional to national and Hubei Province data, complete provincial-level data were publicly available for 22 of 30 remaining provinces: Anhui, Chongqing, Gansu, Guangxi, Guizhou, Hainan, Hebei, Heilongjiang, Henan, Hunan, Inner Mongolia, Jiangsu, Jiangxi, Jilin, Liaoning, Qinghai, Shaanxi, Shandong, Shanxi, Tianjin, Tibet, and Zhejiang. Eight provinces, comprising 26% of the total population, were excluded due to no or insufficient reported data (Beijing, Fujian, Guangdong, Ningxia, Shanghai, Sichuan, Xinjiang, and Yunnan). To understand broader trends despite missing data from the eight excluded provinces, data for Hubei Province were compared with all other provinces combined by calculating the difference between national totals and totals for Hubei Province.

## Results

During epidemiologic weeks 4–9, NHC reported 80,968 new COVID-19 cases (Hubei Province = 67,608 [83% of total new cases reported]), and 659,899 contacts traced (Hubei Province = 264,878 [40% of total contacts traced]) (Table). Reports occasionally noted slight corrections to reported new cases without specifying dates. During the same time period, the 22 included non-Hubei provinces reported an aggregate total of 9,664 cases and 306,684 contacts traced. Among these 22 provinces (Table), those with the largest number of reported cases and contacts traced were Henan (reported cases = 1,274/9,664 [13%]; reported contacts = 39,199/306,684 [13%]) and Zhejiang (reported cases = 1,216/9,664 [13%]; reported contacts = 41,050/306,684 [13%]).

**Table.**
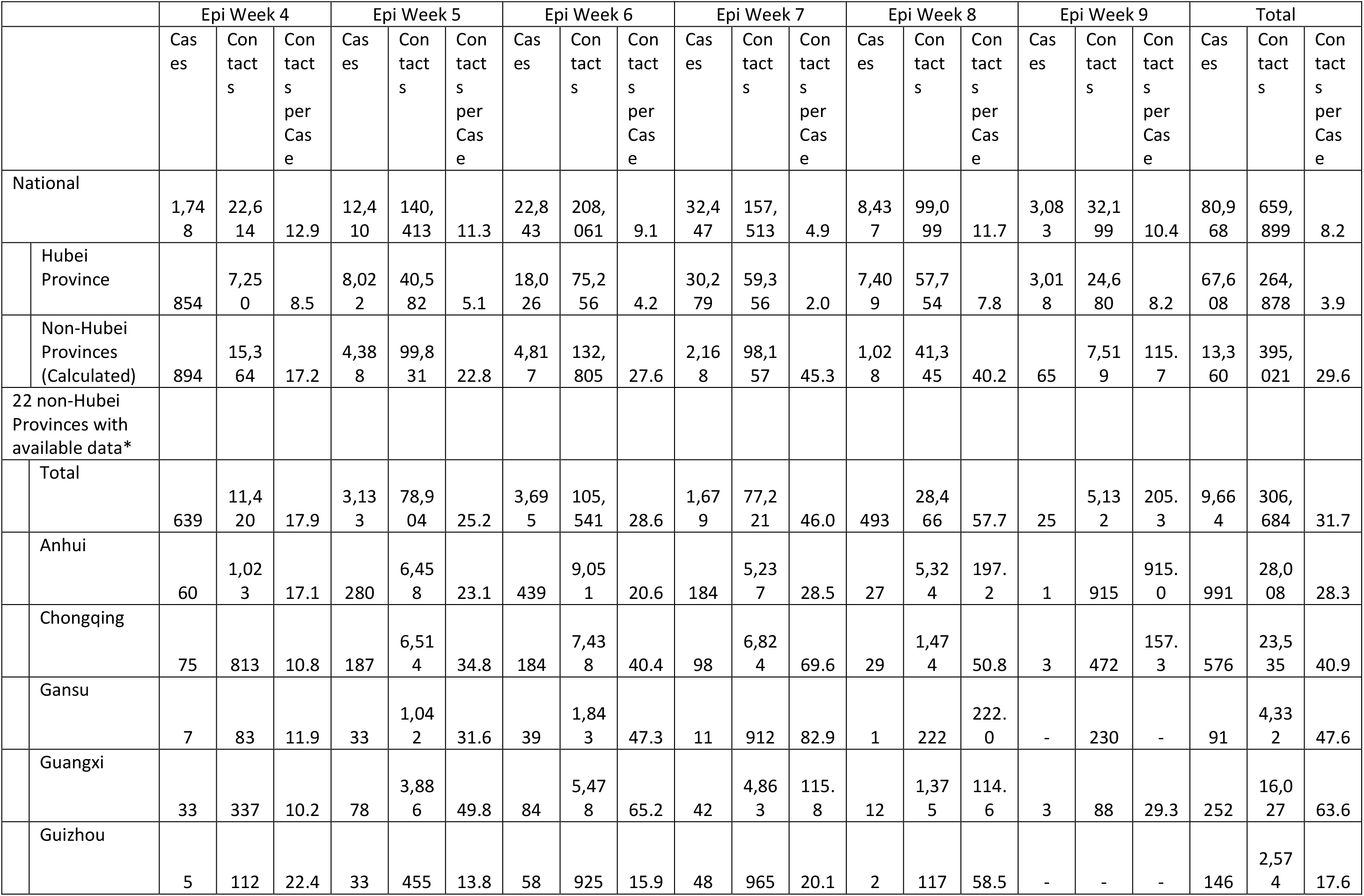
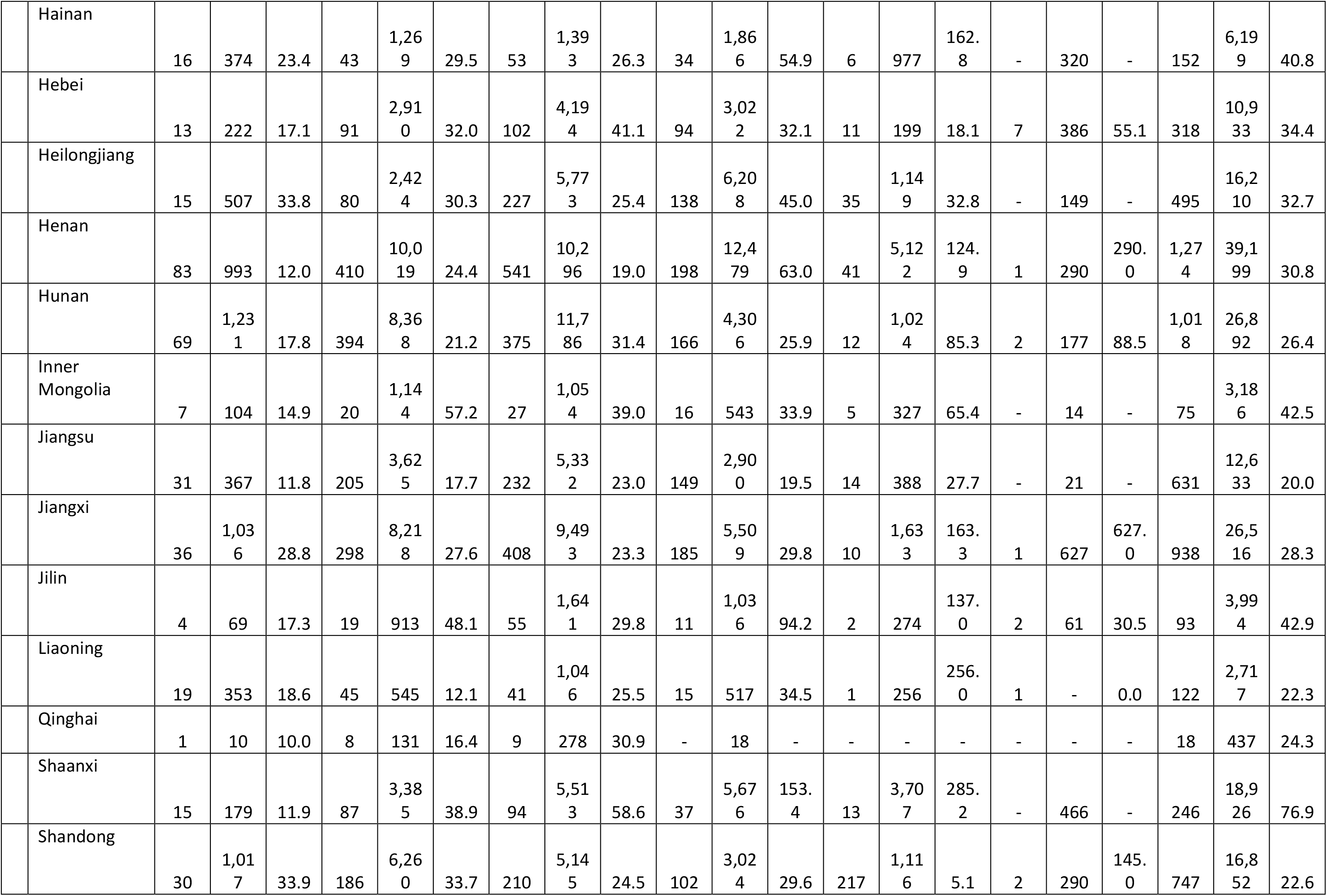
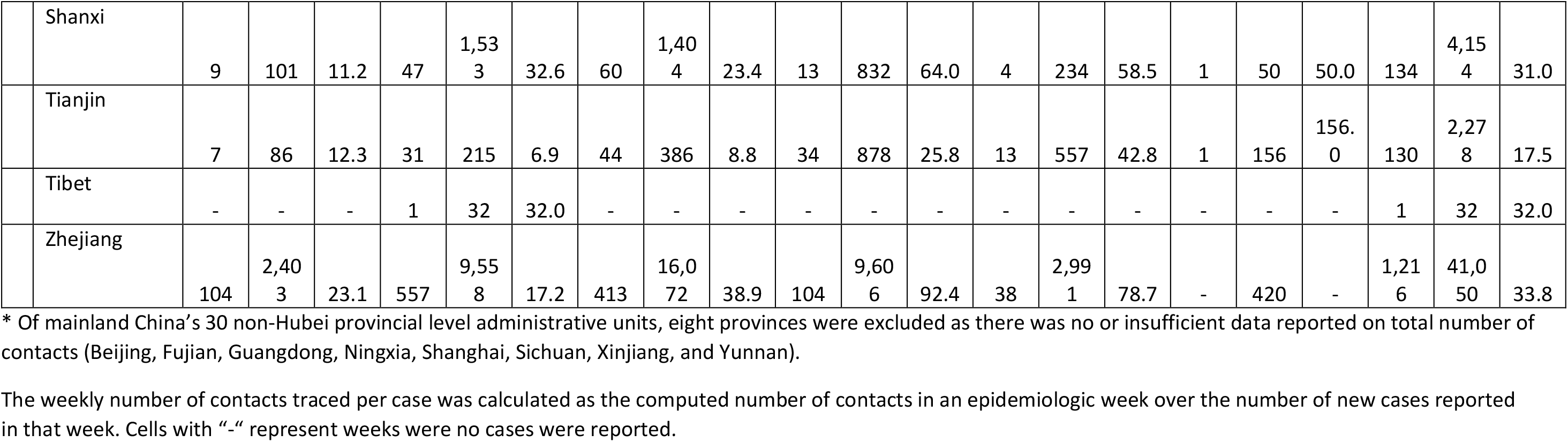
Weekly number of reported Covid-19 cases, contacts traced, and contacts traced per Covid-19 case, by geographic unit, epidemiologic week 4–9, 2020.

|  |  | Epi Week 4 |  |  | Epi Week 5 |  |  | Epi Week 6 |  |  | Epi Week 7 |  |  | Epi Week 8 |  |  | Epi Week 9 |  |  | Total |  |  |
| --- | --- | --- | --- | --- | --- | --- | --- | --- | --- | --- | --- | --- | --- | --- | --- | --- | --- | --- | --- | --- | --- | --- |
|  |  | Cas<br>es | Con<br>tact<br>s | Con<br>tact<br>s<br>per<br>Cas<br>e | Cas<br>es | Con<br>tact<br>s | Con<br>tact<br>s<br>per<br>Cas<br>e | Cas<br>es | Con<br>tact<br>s | Con<br>tact<br>s<br>per<br>Cas<br>e | Cas<br>es | Con<br>tact<br>s | Con<br>tact<br>s<br>per<br>Cas<br>e | Cas<br>es | Con<br>tact<br>s | Con<br>tact<br>s<br>per<br>Cas<br>e | Cas<br>es | Con<br>tact<br>s | Con<br>tact<br>s<br>per<br>Cas<br>e | Cas<br>es | Con<br>tact<br>s | Con<br>tact<br>s<br>per<br>Cas<br>e |
| National |  | 1,748 | 22,614 | 12.9 | 12,410 | 140,413 | 11.3 | 22,843 | 208,061 | 9.1 | 32,447 | 157,513 | 4.9 | 8,437 | 99,099 | 11.7 | 3,083 | 32,199 | 10.4 | 80,968 | 659,899 | 8.2 |
|  | Hubei Province | 854 | 7,250 | 8.5 | 8,022 | 40,582 | 5.1 | 18,026 | 75,256 | 4.2 | 30,279 | 59,356 | 2.0 | 7,409 | 57,754 | 7.8 | 3,018 | 24,680 | 8.2 | 67,608 | 264,878 | 3.9 |
|  | Non-Hubei Provinces (Calculated) | 894 | 15,364 | 17.2 | 4,388 | 99,831 | 22.8 | 4,817 | 132,805 | 27.6 | 2,168 | 98,157 | 45.3 | 1,028 | 41,345 | 40.2 | 65 | 7,519 | 115.7 | 13,360 | 395,021 | 29.6 |
| 22 non-Hubei Provinces with available data* |  |  |  |  |  |  |  |  |  |  |  |  |  |  |  |  |  |  |  |  |  |  |
|  | Total | 639 | 11,420 | 17.9 | 3,133 | 78,904 | 25.2 | 3,695 | 105,541 | 28.6 | 1,679 | 77,221 | 46.0 | 493 | 28,466 | 57.7 | 25 | 5,132 | 205.3 | 9,664 | 306,684 | 31.7 |
|  | Anhui | 60 | 1,023 | 17.1 | 280 | 6,458 | 23.1 | 439 | 9,051 | 20.6 | 184 | 5,237 | 28.5 | 27 | 5,324 | 197.2 | 1 | 915 | 915.0 | 991 | 28,008 | 28.3 |
|  | Chongqing | 75 | 813 | 10.8 | 187 | 6,514 | 34.8 | 184 | 7,438 | 40.4 | 98 | 6,824 | 69.6 | 29 | 1,474 | 50.8 | 3 | 472 | 157.3 | 576 | 23,535 | 40.9 |
|  | Gansu | 7 | 83 | 11.9 | 33 | 1,042 | 31.6 | 39 | 1,843 | 47.3 | 11 | 912 | 82.9 | 1 | 222 | 222.0 | - | 230 | - | 91 | 4,332 | 47.6 |
|  | Guangxi | 33 | 337 | 10.2 | 78 | 3,886 | 49.8 | 84 | 5,478 | 65.2 | 42 | 4,863 | 115.8 | 12 | 1,375 | 114.6 | 3 | 88 | 29.3 | 252 | 16,027 | 63.6 |
|  | Guizhou | 5 | 112 | 22.4 | 33 | 455 | 13.8 | 58 | 925 | 15.9 | 48 | 965 | 20.1 | 2 | 117 | 58.5 | - | - | - | 146 | 2,574 | 17.6 |

|  |  |  |  |  |  |  |  |  |  |  |  |  |  |  |  |  |  |  |  |  |  |
| --- | --- | --- | --- | --- | --- | --- | --- | --- | --- | --- | --- | --- | --- | --- | --- | --- | --- | --- | --- | --- | --- |
| Hainan | 16 | 374 | 23.4 | 43 | 1,269 | 29.5 | 53 | 1,393 | 26.3 | 34 | 1,866 | 54.9 | 6 | 977 | 162.8 | - | 320 | - | 152 | 6,199 | 40.8 |
| Hebei | 13 | 222 | 17.1 | 91 | 2,910 | 32.0 | 102 | 4,194 | 41.1 | 94 | 3,022 | 32.1 | 11 | 199 | 18.1 | 7 | 386 | 55.1 | 318 | 10,933 | 34.4 |
| Heilongjiang | 15 | 507 | 33.8 | 80 | 2,424 | 30.3 | 227 | 5,773 | 25.4 | 138 | 6,208 | 45.0 | 35 | 1,149 | 32.8 | - | 149 | - | 495 | 16,210 | 32.7 |
| Henan | 83 | 993 | 12.0 | 410 | 10,019 | 24.4 | 541 | 10,296 | 19.0 | 198 | 12,479 | 63.0 | 41 | 5,122 | 124.9 | 1 | 290 | 290.0 | 1,274 | 39,199 | 30.8 |
| Hunan | 69 | 1,231 | 17.8 | 394 | 8,368 | 21.2 | 375 | 11,786 | 31.4 | 166 | 4,306 | 25.9 | 12 | 1,024 | 85.3 | 2 | 177 | 88.5 | 1,018 | 26,892 | 26.4 |
| Inner Mongolia | 7 | 104 | 14.9 | 20 | 1,144 | 57.2 | 27 | 1,054 | 39.0 | 16 | 543 | 33.9 | 5 | 327 | 65.4 | - | 14 | - | 75 | 3,186 | 42.5 |
| Jiangsu | 31 | 367 | 11.8 | 205 | 3,625 | 17.7 | 232 | 5,332 | 23.0 | 149 | 2,900 | 19.5 | 14 | 388 | 27.7 | - | 21 | - | 631 | 12,633 | 20.0 |
| Jiangxi | 36 | 1,036 | 28.8 | 298 | 8,218 | 27.6 | 408 | 9,493 | 23.3 | 185 | 5,509 | 29.8 | 10 | 1,633 | 163.3 | 1 | 627 | 627.0 | 938 | 26,516 | 28.3 |
| Jilin | 4 | 69 | 17.3 | 19 | 913 | 48.1 | 55 | 1,641 | 29.8 | 11 | 1,036 | 94.2 | 2 | 274 | 137.0 | 2 | 61 | 30.5 | 93 | 3,994 | 42.9 |
| Liaoning | 19 | 353 | 18.6 | 45 | 545 | 12.1 | 41 | 1,046 | 25.5 | 15 | 517 | 34.5 | 1 | 256 | 256.0 | 1 | - | 0.0 | 122 | 2,717 | 22.3 |
| Qinghai | 1 | 10 | 10.0 | 8 | 131 | 16.4 | 9 | 278 | 30.9 | - | 18 | - | - | - | - | - | - | - | 18 | 437 | 24.3 |
| Shaanxi | 15 | 179 | 11.9 | 87 | 3,385 | 38.9 | 94 | 5,513 | 58.6 | 37 | 5,676 | 153.4 | 13 | 3,707 | 285.2 | - | 466 | - | 246 | 18,926 | 76.9 |
| Shandong | 30 | 1,017 | 33.9 | 186 | 6,260 | 33.7 | 210 | 5,145 | 24.5 | 102 | 3,024 | 29.6 | 217 | 1,116 | 5.1 | 2 | 290 | 145.0 | 747 | 16,852 | 22.6 |
| Shanxi | 9 | 101 | 11.2 | 47 | 1,533 | 32.6 | 60 | 1,404 | 23.4 | 13 | 832 | 64.0 | 4 | 234 | 58.5 | 1 | 50 | 50.0 | 134 | 4,154 | 31.0 |
| Tianjin | 7 | 86 | 12.3 | 31 | 215 | 6.9 | 44 | 386 | 8.8 | 34 | 878 | 25.8 | 13 | 557 | 42.8 | 1 | 156 | 156.0 | 130 | 2,278 | 17.5 |
| Tibet | - | - | - | 1 | 32 | 32.0 | - | - | - | - | - | - | - | - | - | - | - | - | 1 | 32 | 32.0 |
| Zhejiang | 104 | 2,403 | 23.1 | 557 | 9,558 | 17.2 | 413 | 16,072 | 38.9 | 104 | 9,606 | 92.4 | 38 | 2,991 | 78.7 | - | 420 | - | 1,216 | 41,050 | 33.8 |

\* Of mainland China's 30 non-Hubei provincial level administrative units, eight provinces were excluded as there was no or insufficient data reported on total number of contacts (Beijing, Fujian, Guangdong, Ningxia, Shanghai, Sichuan, Xinjiang, and Yunnan).
The weekly number of contacts traced per case was calculated as the computed number of contacts in an epidemiologic week over the number of new cases reported in that week. Cells with “-” represent weeks where no cases were reported.

The weekly number of contacts traced per case remained below 10 in Hubei Province (range = 2.0 in epidemiologic week 7 to 8.2 in epidemiologic week 4); the lowest value in epidemiologic week 7 occurred when 18,453 clinically diagnosed cases were included in reported case counts for February 12–15, increasing the denominator substantially (Figure 1). By comparison, in non-Hubei provinces the weekly number of contacts traced per case was higher than in Hubei Province and increased from 17.2 in epidemiologic week 4 to 115.7 in epidemiologic week 9. Provincial data from the 22 included non-Hubei provinces indicate that the number of contacts traced per case increased as case counts declined while reported number of contacts traced remained high or increased (Table). For example, Anhui Province reported 60 cases with 1,023 reported contacts traced during epidemiologic week 4 (17.1 contacts traced per case) compared to 1 case with 915 reported contacts traced during epidemiologic week 9 (915 contacts traced per case).

**Figure.**
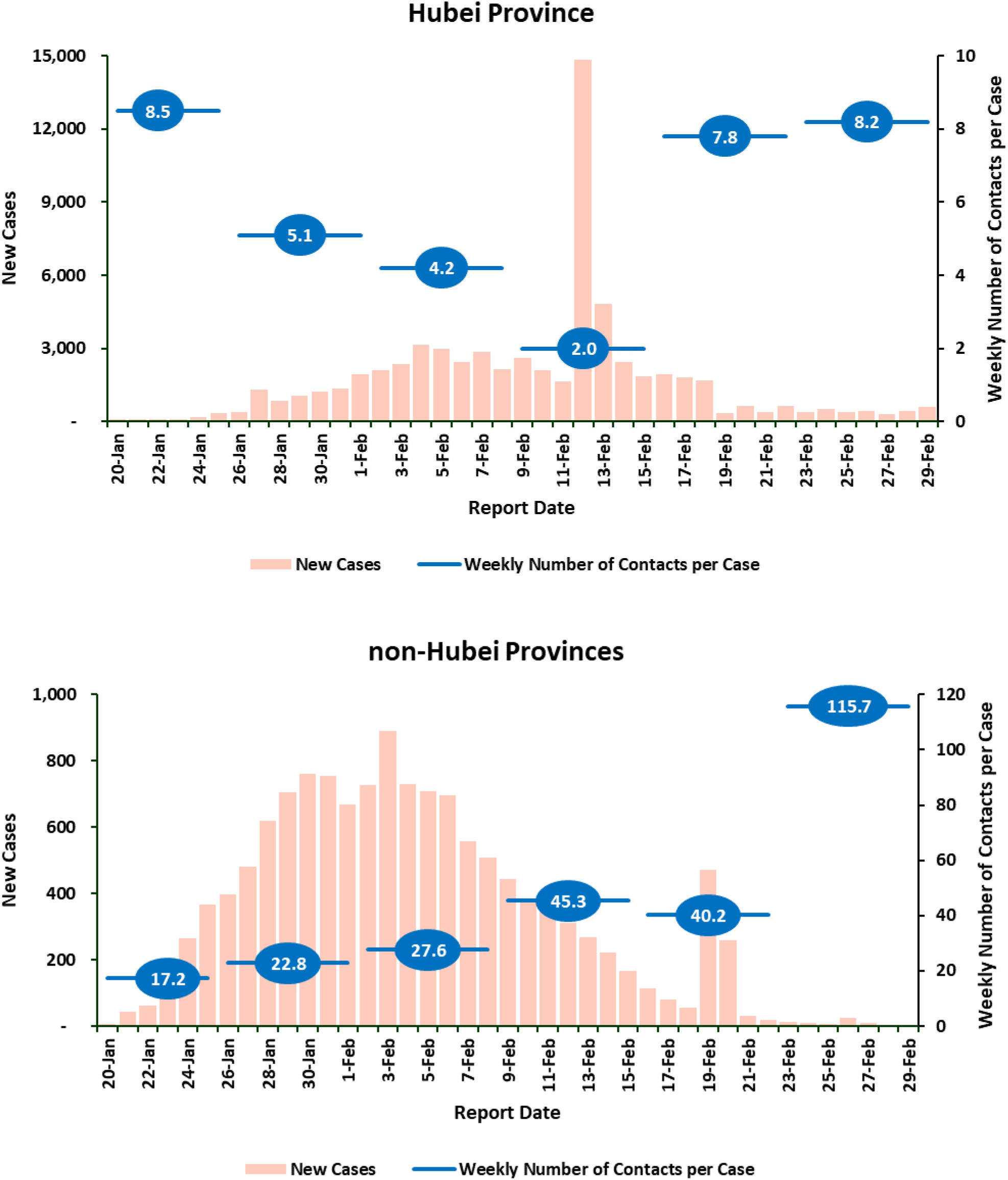
Reported COVID-19 cases and weekly number of contacts traced per case, by week, Hubei Province vs non-Hubei Provinces, epidemiologic weeks 4–9. Weekly number of contacts per case was calculated as the computed number of new contacts in an epidemiologic week divided by the number of new cases reported in that week. In Hubei Province, the lowest value in epidemiologic week 7 occurred when 18,453 clinically diagnosed cases were included in reported case counts for February 12–15, increasing the denominator substantially.

## Discussion

Along with other NPI used in China, contact tracing likely contributed to reducing SARS-CoV-2 transmission by quarantining a large number of potentially infected contacts (9). It potentially helped to identify pre-symptomatic and asymptomatic infections early and reduced the time from symptom onset to initiation of medical care (13, 14). Our analysis suggests that contact tracing implementation and data reporting varied by province, with non-Hubei provinces reporting similar trends that contrast trends in Hubei Province. Notably, despite reporting only 15% of total cases, non-Hubei provinces had 1.5 times more reported contacts traced compared to Hubei Province. Provincial-level differences likely reflect local capacity for implementing contact tracing, differences in local disease transmission, and evolving guidelines. These results suggest that contract tracing may have been more complete in areas and periods with lower case counts, which was potentially linked to resource constraints.

Contact tracing with quarantine is resource intensive. For example, in Wuhan City, contact tracing was conducted by 1,800 epidemiologists working in teams of five (8). Data on provincial level contact tracing resources were not available. Geographic and temporal differences may reflect variability in available resources, including trained staff for contact tracing and medical observation, housing for contacts, and laboratory testing capacity. While contact tracing identified and isolated a large number of potentially infected contacts, published studies showed most contacts did not become reported cases; 30.4% (391 positive contacts/1,286 contacts traced) in Shenzhen, 2.6% (129/4,950) in Guangzhou, and 2.3% (120 /5,241) in Xi’an (13-15).

This report has several limitations. First, without individual patient-level data, our analysis is based on aggregate data and subject to ecological fallacy. Second, data were compiled from publicly available online reports, and we did not have access to primary data. This required excluding eight provinces and did not allow analysis of available contact tracing resources. As such, data could not be externally verified, and the data collection methods were not available, including confirmation that all reported contacts traced were linked to reported confirmed cases. Second, inter-provincial variability may have affected reported data comparability, and eight provinces were excluded. Finally, we do not know the distribution of contacts for individual cases. The actual number of contacts traced likely differed by exposure type (e.g., family, shopping center, public transport), and the proxy (mean contacts traced per case) would overestimate median contacts per case when large numbers of contacts were linked to a single case (i.e., attending a public gathering with a confirmed case).

Despite these limitations, our findings help describe contact tracing in China as part of the COVID-19 response. Future investigations can aim to better understand the role of COVID-19 contact tracing and quarantine, including timeliness of contact tracing and quarantine, prioritization of contacts who are more likely associated with viral transmission and the effectiveness of contact tracing in differing epidemiologic, social, and resource availability contexts.

## Data Availability

Data were compiled from Provincial-Level Health Commission Websites Containing Publicly Available Reported Data on COVID-19
National Health Commission http://weekly.chinacdc.cn/news/TrackingtheEpidemic.htm
Anhui http://wjw.ah.gov.cn/
Beijing http://wjw.beijing.gov.cn/xwzx_20031/xwfb/
Chongqing http://wsjkw.cq.gov.cn/
Fujian http://wjw.fujian.gov.cn/
Gansu http://wsjk.gansu.gov.cn/
Guangdong http://wsjkw.gd.gov.cn/zwyw_yqxx/index.html
Guangxi http://wsjkw.gxzf.gov.cn/gzdt/bt/
Guizhou http://www.gzhfpc.gov.cn/
Hainan http://wst.hainan.gov.cn/swjw/index.html
Hebei http://wsjkw.hebei.gov.cn/
Heilongjiang http://wsjkw.hlj.gov.cn/
Henan http://www.hnwsjsw.gov.cn/
Hubei http://wjw.hubei.gov.cn/fbjd/dtyw/
Hunan http://wjw.hunan.gov.cn/
Inner Mongolia http://wjw.nmg.gov.cn/
Jiangsu http://wjw.jiangsu.gov.cn/
Jiangxi http://hc.jiangxi.gov.cn/
Jilin http://wsjkw.jl.gov.cn/
Liaoning http://wsjk.ln.gov.cn/
Ningxia http://wsjkw.nx.gov.cn/
Qinghai https://wsjkw.qinghai.gov.cn/
Shaanxi http://sxwjw.shaanxi.gov.cn/
Shandong http://wsjkw.shandong.gov.cn
Shanghai http://wsjkw.sh.gov.cn/xwfb/index.html
Shanxi http://wjw.shanxi.gov.cn/
Sichuan http://wsjkw.sc.gov.cn/scwsjkw/szyw/tygl.shtml
Tianjin http://wsjs.tj.gov.cn/
Tibet http://wjw.xizang.gov.cn
Xinjiang http://xjhfpc.gov.cn
Yunnan http://ynswsjkw.yn.gov.cn/wjwWebsite/web/index
Zhejiang http://www.zjwjw.gov.cn/col/col1202101/index.html

## Technical Appendix

Provincial-Level Health Commission Websites Containing Publicly Available Reported Data on COVID-19

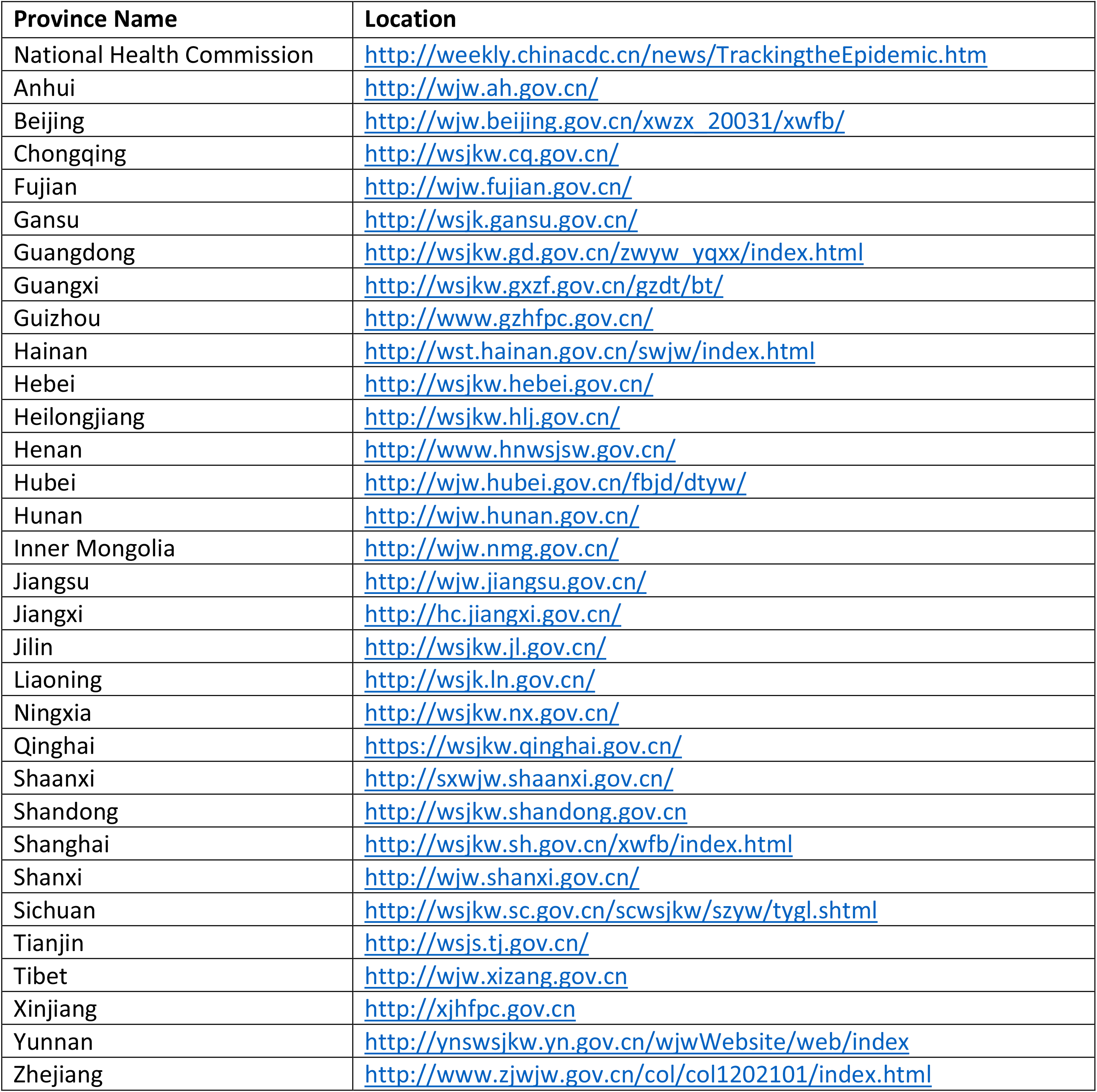

## References

1. Pan A, Liu L, Wang C, Guo H, Hao X, Wang Q, et al. Association of Public Health Interventions with the Epidemiology of the COVID-19 Outbreak in Wuhan, China. JAMA. 2020;323(19):1915–23. doi: 10.1001/jama.2020.6130.

2. China National Health Commission. Tracking the Epidemic. 2020 [cited May 7, 2020]; Available from: http://weekly.chinacdc.cn/news/TrackingtheEpidemic.htm

3. Qualls N, Levitt A, Kanade N, Wright-Jegede N, Dopson S, Biggerstaff M, et al. Community Mitigation Guidelines to Prevent Pandemic Influenza — United States, 2017. MMWR Recomm Rep. 2017 Apr 21;66(1):1–34. doi: 10.15585/mmwr.rr6601a1.

4. Fong MW, Gao H, Wong J, Y., Xiao J, Shiu EYC, Ryu S, et al. Nonpharmaceutical Measures for Pandemic Influenza in Nonhealthcare Settings — Social Distancing Measures. Emerg Infect Dis. 2020;26(5):976–84. doi: 10.3201/eid2605.190995.

5. Cheng H, Jian S, Liu D, Ng T, Huang W, Lin H, et al. Contact Tracing Assessment of COVID-19 Transmission Dynamics in Taiwan and Risk at Different Exposure Periods Before and After Symptom Onset. JAMA Intern Med. 2020:E1–E8. doi: 10.1001/jamainternmed.2020.2020.

6. Ng Y, Li Z, Chua Y, Chaw W, Zhao Z, Er B, et al. Evaluation of the Effectiveness of Surveillance and Containment Measures for the First 100 Patients with COVID-19 in Singapore — January 2–February 29, 2020. MMWR Morb Mortal Wkly Rep. 2020 Mar 20;69(11):307–11.

7. Covid-19 National Emergency Response Center E, Case Management Team KCDC. Coronavirus Disease-19: Summary of 2,370 Contact Investigations of the First 30 Cases in the Republic of Korea. Osong Public Health Res Perspect. 2020;11(2):81–4. doi: 10.24171/j.phrp.2020.11.2.04.

8. WHO-China Joint Mission. Report of the WHO-China Joint Mission on Coronavirus Disease 2019 (COVID-19). Geneva: WHO; 2020.

9. Lai S, Ruktanonchai NW, Zhou L, Prosper O, Luo W, Floyd JR, et al. Effect of non-pharmaceutical interventions to contain COVID-19 in China. Nature. 2020 2020/05/04.

10. Novel Coronavirus Pneumonia Emergency Response Epidemiology Team. The Epidemiological Characteristics of an Outbreak of 2019 Novel Coronavirus Diseases (COVID-19) — China, 2020. China CDC Weekly. 2020;2(8):113–22. doi: 10.1038/s41586-020-2293-x.

11. Chinese Center for Disease Control and Prevention. Guidelines for COVID-19 Epidemiological Investigations. China CDC Weekly. 2020;2(19):327–8.

12. Chinese Center for Disease Control and Prevention. Guideline for Investigation and Management of Close Contacts of COVID-19 Cases. China CDC Weekly. 2020;2(19):329–31.

13. Bi Q, Wu Y, Mei S, Ye C, Zou X, Zhang Z, et al. Epidemiology and transmission of COVID-19 in 391 cases and 1286 of their close contacts in Shenzhen, China: a retrospective cohort study. Lancet Infect Dis. 2020:1–9. doi: 10.1016/S1473-3099(20)30287-5.

14. Luo L, Liu D, Liao X, Wu X, Jing Q, Zheng J, et al. Modes of contact and risk of transmission in COVID-19 among close contacts. medRxiv. 2020. doi: https://doi.org/10.1101/2020.03.24.20042606

15. Zhang H, Ji Z, CHeng Z, Zeng L, Mi B, Cheng F, et al. [Epidemiological characteristics of close contact in Xi’an]. J of Xi’an Jiaotong University (Medical Sciences). 2020:1–7. ISSN:1671-8259/CN:61-1399/R

